# Genome-wide association study of a semicontinuous trait: Illustration of the impact of the modeling strategy through the study of Neutrophil Extracellular Traps levels

**DOI:** 10.1101/2022.09.19.22279929

**Authors:** Gaëlle Munsch, Carole Proust, Sylvie Labrouche-Colomer, Dylan Aïssi, Anne Boland, Pierre-Emmanuel Morange, Anne Roche, Luc de Chaisemartin, Annie Harroche, Robert Olaso, Jean-François Deleuze, Chloé James, Joseph Emmerich, David M Smadja, Hélène Jacqmin-Gadda, David-Alexandre Trégouët

**Affiliations:** Univ. Bordeaux, Inserm, Bordeaux Population Health Research Center, UMR 1219, F-33000 Bordeaux, France; UMR1034, Inserm, Biology of Cardiovascular Diseases, University of Bordeaux, Pessac, France; Laboratoire d’Hématologie, CHU de Bordeaux, Pessac, France; Université Paris-Saclay, CEA, Centre National de Recherche en Génomique Humaine (CNRGH), 91057, Evry, France; INSERM UMR_S 1263, Nutrition Obesity and Risk of Thrombosis, Center for CardioVascular and Nutrition research (C2VN), Aix-Marseille University, Marseille 13385, France; Laboratory of Haematology, La Timone Hospital, Marseille 13385, France; Service pneumologie hôpital Bicêtre; Service Auto-immunité, Hypersensibilité et Biothérapies, Hôpital Bichat, Assistance Publique-Hôpitaux de Paris, Paris, France; Université Paris-Saclay, INSERM, Inflammation, Microbiome, Immunosurveillance, Orsay, France; Service d’Hématologie Clinique Centre de Traitement de l’Hémophilie Hôpital Necker Enfants Malades; Centre d’Etude du Polymorphisme Humain, Fondation Jean Dausset, Paris, France; Department of vascular medicine, Paris Saint-Joseph Hospital Group, University of Paris, 185145 rue Raymond Losserand, Paris, 75674, France; UMR1153, INSERM CRESS, 185 rue Raymond Losserand, Paris, 75674, France; Innovative Therapies in Hemostasis, Université de Paris, INSERM, F-75006, Paris, France; Hematology Department and Biosurgical Research Lab (Carpentier Foundation), Assistance Publique Hôpitaux de Paris, Centre-Université de Paris (APHP-CUP), F-75015, Paris, France

**Keywords:** Semicontinuous outcome, Compound Poisson-Gamma, Negative Binomial, Genome Wide Association Study, Neutrophil Extracellular Traps

## Abstract

Semicontinuous data, characterized by an excess of zeros followed by a non-negative and right-skewed distribution, are frequently observed in biomedical research. Different statistical models have been proposed to investigate the association of covariates with such outcome. Motivated by the search of genetic factors associated with Neutrophil Extracellular Traps (NETs), a semicontinuous biomarker involved in thrombosis, we here investigated the impact of the selected model for semicontinuous traits in the context of a Genome Wide Association Study (GWAS). We compared three models that jointly model zero and positive values while allowing the estimation of a single association parameter of covariates with the global mean: Tobit, Negative Binomial and Compound Poisson-Gamma. We assessed the fit of these models to a sample of 657 participants of the FARIVE study measured for NETs plasma levels. For each of these three models, we performed a GWAS on NETs in FARIVE participants and results were compared. A simulation study was also conducted to evaluate the control of the type I error. Compound Poisson-Gamma and Negative Binomial models fitted NETs data observed in FARIVE better than the Tobit model. However, the Negative Binomial model suffered from an inflation of its type I error, attributable to extreme positive values of the NETs and low frequency variants. Conversely, the Compound Poisson-Gamma model was robust to both phenomena. Using the latter model, a GWAS identified a genome wide significant locus on chr21q21.3. The lead variant was rs57502213, a deletion of two nucleotides located ∼40kb upstream the non-coding RNA (miR155HG) hosting the miR-155 that was recently highlighted to have a role in NETs formation. This work indicates that the modeling strategy for a semicontinuous outcome in the framework of GWAS studies is crucial. The choice of the model should take into account the nature of the process generating zero values and the presence of extreme values. Our work also suggests that the Compound Poisson-Gamma model, while still marginally employed, can be a robust modeling strategy for GWAS analysis on a semicontinuous trait.

## Introduction

Semicontinuous data, characterized by an excess of zeros followed by a non-negative and right-skewed distribution, are frequently observed in biomedical research^1^. When the study aims at identifying determinants of such a semicontinuous biomarker, it must be handled as the outcome variable and due to the inflation of zeros, classical models such as linear regression cannot be applied without violating the Gaussian assumptions, even with a logarithmic or rank-based inverse-normal transformation. For instance when the interest specifically lies in the identification of molecular determinants associated with a disease semicontinuous biomarker, as it is encountered in the omics era in order to identify/characterize new biological pathways, inform about drug discovery and help in individual risk prediction^2^, the problem of how to model its distribution arises.

Over the past decades several statistical models have been developed to model semicontinuous data by taking into account the mass of zeros. Among the most commonly used models are the Tobit and the two-part models^3,4^.

The two-part model and its extensions^5,6^ rely on the use of a logistic regression model to predict the probabilities of occurrence of zero values and of a linear regression model for the analysis of the strictly continuous outcome. The main assumption of this model is that the values of the outcome are derived from two different generating processes. This model has been used in various applications including the modeling of tumor size in cancer, food intake, microbiome abundance or individual costs of chronic kidney disease^7–11^. The two-part model, does not make possible the estimation of a single parameter that represents the association of an explanatory variable on the outcome. In contrast to the two-part model, Tobit models consider a single distribution of the outcome. In the case of zero-inflated data, the Tobit model assumes that the semicontinuous variable is a truncated observation of a Gaussian variable. This modeling is mainly used to account for floor or ceiling effect of the outcome variable that could be due to technical measurement limits^12–15^.

Another possibility is to consider the outcome variable as quantitative discrete, which can be done in some cases by changing the unit of measurement through the use of a multiplicative factor, without losing precision. In this case, models for count data such as the Poisson model or the Negative Binomial model in presence of overdispersion can be used. These models are relevant as long as the proportion of zeros is not too high^16,17^. As the Tobit model, these models allow for a simple interpretation of the results since only one coefficient is estimated per explanatory variable. Extensions of these models have been developed to account for the zero mass (also known as ZIP for Zero-Inflated Poisson and ZINB for Zero-Inflated Negative Binomial) but they make the assumption that the distribution of the outcome is composed of two generating processes, like the two-part models.

New models based on so-called Tweedie distributions^18–20^ have recently emerged for the analysis of semicontinuous data but their use remains marginal^21^. The Compound Poisson-Gamma model belongs to this Tweedie family. It assumes that the semicontinuous outcome is defined as a Poisson sum of gamma random variables. Semicontinuous data are then modeled through the use of a single distribution.

The choice between these different models is not obvious as each semicontinuous outcome has its own properties. As there is no established decision tool, the model to be applied should be chosen according to the distribution of the outcome and the clinical context^22^.

In this work, we show the impact of the adopted model on the results of a Genome Wide Association Study (GWAS) that aimed at identifying genetic factors associated with plasma levels of Neutrophil Extracellular Traps (NETs), a semicontinuous biomarker involved in thrombosis. We highlight the differences between the models with respect to the flexibility of the underlying assumptions, the robustness to outliers and low allele frequency that can help to select the most appropriate model for future studies.

NETs are one of the emerging biomarkers with a key role in thrombosis^23^. In the event of a vascular breach, neutrophils and platelets are the first cells to be recruited and activated^24^. When neutrophils are activated by platelets, they have pro-inflammatory properties that can enhance tissue damage and induce thrombus formation in particular when they evolve towards a certain form of cell death leading to the release of their decondensed chromatin as a network of fibres also called NETs. NETs are composed of DNA fibres comprised of antimicrobial proteins and histones which promote coagulation, platelets activation and thus thrombus formation^25,26^. NETs are involved in many other biological mechanisms such as immune response to viruses, diabetes, cystic fibrosis, cancer tumor growth, progression and metastasis^27–31^.

NETs plasma levels were here measured in 657 participants of the « FActeurs de RIsque et de récidives de la maladie thromboembolique Veineuse » (FARIVE) study^32^. Genome wide genotype data were also available for these participants and then used to conduct a GWAS on NETs levels. We illustrate how the results of this GWAS are impacted by the statistical approach adopted to model NETs plasma levels.

## Methods

### The FARIVE study

The FARIVE study is a multicenter case-control study conducted between 2003 and 2007. The sample includes 607 patients with a documented episode of deep vein thrombosis and/or pulmonary embolism and 607 healthy individuals. A detailed description of the study can be found elsewhere^32^. Briefly, patients were not eligible if they were younger than 18 years, had previous venous thrombosis (VT) event, active cancer or recent history of malignancy (within 5 years). Controls were recruited over the same period and matched to cases according to age and sex. They did not have any history of venous and arterial thrombotic disease as well as cancer, liver or kidney failure.

#### 1. NETs measurements

Neutrophil Extracellular Traps (NETs) were quantified by measuring myeloperoxidase (MPO)-DNA complexes using an in-house capture ELISA already described^33^ in a subsample of 410 VT patients (7 months after their inclusion in the study once the anticoagulant treatment has stopped) and 327 controls (at their time of inclusion in the study). Briefly, microtiter plates were coated with anti-human MPO antibody. After blocking, serum samples were added together with a peroxidase-labeled anti-DNA antibody. After incubation, the peroxidase substrate was added and absorbance measured at 405 nm in a spectrophotometer.

#### 2. Genotyping and Imputation

FARIVE participants were genotyped using the Illumina Infinium Global Screening Array v3.0 (GSAv3.0) microarray at the Centre National de Recherche en Génomique Humaine (CNRGH). Individuals with at least one of the following criteria were excluded: discordant sex information, relatedness individuals identified by pairwise clustering of identity by state distances (IBS), genotyping call rate lower than 99%, heterozygosity rate higher/lower than the average rate ± 3 standard deviation or of non-European ancestry. After applying these criteria, the final sample was composed of 1,077 individuals. Among the 730,059 variants genotyped, 145,238 variants without a valid annotation were excluded as well as 656 variants deviating from Hardy-Weinberg equilibrium in controls at *P*<10^-6^, 47,286 variants with a Minor Allele Count (MAC) lower than 20 and 1,774 variants with a call rate lower than 95%. This quality controls procedure was conducted using Plink v1.9^34^ and the R software v3.6.2. Finally, there were 535,105 markers left for imputation which was then performed with Minimac4 using the 1000 Genomes phase 3 version 5 reference panel^35^.

#### 3. Genome Wide Association Study of NETs plasma levels

The present study relies on a subsample of 657 individuals (372 VT cases and 285 controls) with both NETs measurements and imputed genetic data. All single nucleotide polymorphisms (SNPs) with minor allele frequency (MAF) greater than 0.01 and imputation quality score greater than 0.3 were tested for association with NETs plasma levels. As shown in the next section, 3 different statistical models were deployed. In all, associations were tested on imputed allelic doses and adjusted for potential confounders that is age, sex, smoking, case-control status and the four first principal components derived from genome wide genotype data^36–38^. The standard genome-wide statistical threshold of 5×10^-8^ was used to consider SNPs as significantly associated with NETs plasma levels.

### Statistical modeling for GWAS analysis of NETs plasma levels

Since we were interested in identifying genetic factors that influence mean NETs plasma levels, any statistical approach that treats independently the zeros mass and the distribution of positive values, such as the two-part, ZIP and ZINB models, was deemed not adapted to our application. As a consequence, only three models were compared in this study: the Tobit, Compound Poisson-Gamma, and Negative Binomial models. The Poisson model was not investigated because NETs plasma data presented a large overdispersion (see Results section), a situation where Negative Binomial model is preferable.

In this study, we aimed to identify the most suitable model for semicontinuous data in order to conduct a GWAS on NETs and highlight the difference between the three models.

#### 1. Tobit model

In the Tobit model, the observed variable Y is assumed to be a right or left truncated observation of an underlying Gaussian latent variable (*Y*\*). Let the constant threshold for truncation which needs to be known and is equal to zero in the context of zero-inflated data. Therefore, the Tobit model assumes that zero values are due to censoring or measurement limits and so they do not represent the true absence of the variable. In the case of a left truncation at 0 the values of the observed variable are:

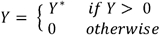

The subsequent regression model is:

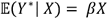

where 𝔼(*Y*\*| *X*) is the expected value of the underlying Gaussian variable *Y*\* conditioned on the explanatory variables *X*, and where *β* represents the regression parameters associated to *X*. The Tobit model is available in the *VGAM* R package^39^.

#### 2. Compound Poisson-Gamma model

An Exponential Dispersion Model (EDM) is a two-parameter family of distributions composed of a linear exponential family with an additional dispersion parameter^40^. EDMs are characterized by their variance function 𝕍(·) that is an exponential function used to describe the relationship between the mean and the variance. If *Y* follows an EDM, then 𝔼(*Y*)= *µ* and *Var* (*Y*) = *Φ* 𝕍(*µ*) with *Φ* a dispersion parameter. Tweedie models are a class of EDMs characterized by a power variance function: 𝕍(*µ*) = *µ*^*p*^ with *p* the index parameter^41,42^. Most of the usual distributions are included in the class of Tweedie models such as the normal (*p* = 0), Poisson (*p* = 1), gamma (*p* = 2) and the inverse Gaussian (*p* = 3)^43^.

The probability density function of a Tweedie model is defined as^40^:

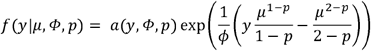

where *a*(·) is a given function.

The Compound Poisson-Gamma model belongs to the family of Tweedie models with *p* ∈] 1;2[. It simultaneously models the occurrence and the intensity of the semicontinuous outcome^44^. The distribution of a variable Y following a Compound Poisson-Gamma model may be defined as a Poisson sum of *M* Gamma distributions:

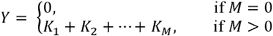

where *M* ∼*Pois*(*λ*), *K*_*i*_ ∼*Gamma*(*α, γ*) with *α* the shape parameter *γ* and the scale parameter, and where the values of *K*_*i*_ are *iid* and independent on *M*.

The Compound Poisson-Gamma model is a Tweedie model with the following parametrisation:

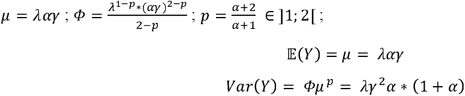

Thus, direct modeling of the global expectation 𝔼(*Y*) is possible using a generalized linear model with a logarithmic link function to insure positivity of the means:

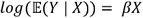

We used the *cplm* R package to implement Compound Poisson-Gamma models^45^.

#### 3. Negative Binomial models

We also attempted to use a model for count data by multiplying NETs’ values by 1,000 to ensure discreteness without creating new ex-aequos. Let be a random variable following a Poisson distribution which depends on a single parameter *λ* > 0:

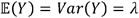

The Poisson model is adapted to model the expectation of a count variable using a generalized linear model with a logarithmic link function:

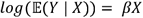

The Negative Binomial model is an extension to the Poisson model in the presence of over-dispersion of the outcome: *Var* (*Y*) > *E*(*Y*)^46,47^. In that case, the variance of Y is linked to its expectation through the following relationship: where *Var* (*Y*) = 𝔼(*Y*) + *k \** 𝔼(*Y*)^2^ where *k* > 0 is a dispersion parameter. This model is also part of generalized linear models and its link function is the logarithm. Negative Binomial model can also be represented as Poisson distributions with a Gamma distributed means where Y ∼*Pois*(*λ*) and *λ* ∼ *Gamma*(*α, γ*)^48^. However, unlike the Compound Poisson-Gamma presented above, the two variables and are not independent from each other.

### Models’ comparison

The three aforementioned tested models were applied to NETs data while adjusting for age, sex, smoking and case-control status.

The fit of these models to NETs data were assessed in two ways. First, we computed the Root Mean Square Error (RMSE) of each tested model defined as:

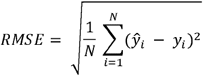

where N is the sample size, *ŷ*_*i*_ the prediction of the *i*^*th*^ individual according to its covariates provided by a given modeling approach and *y*_*i*_ the observed value. Instead of predicting *ŷ*_*i*_ by 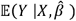 that cannot be equal to zero, we used simulated predictions. For each studied model, a random value was generated for each individual according to its explanatory variables and the estimated model parameters. This process was repeated 1,000 times and the mean of RMSEs over the 1000 replicates was reported. As the Tobit model predicts negative values that are not observed in our semicontinuous outcome, these were imputed at zero to calculate the corresponding RMSE.

Second, we graphically assessed the fit of models predictions using Quantile-Quantile plot (QQplot) of the observations and predictions for each tested model.

### Simulation Study

A simulation study was conducted to evaluate the control of the type I error (*α*) of the Negative Binomial and Compound Poisson-Gamma models in the context of genetic association studies as well as their sensibility to outliers. From the observed NETs data distribution, we randomly generated *S* = 1,…,10000 bootstrapped samples of size *N* = 657. For each bootstrapped sample, all individuals were randomly assigned 4 independent genotypes under the assumption of Hardy-Weinberg and corresponding to 4 SNPs with allele frequencies 0.01, 0.05, 0.10 and 0.20. The association of SNPs with the outcome was tested under the assumption of additive allele effects. This procedure was used to simulate semicontinuous data that mimic the NETs data observed in FARIVE and to allow the evaluation of the robustness of the two studied models (Negative Binomial and Compound Poisson-Gamma models) to a deviation from their underlying distribution. To assess the robustness to outliers, each simulated dataset was also analysed after the exclusion of individuals with simulated trait higher than 0.5, a threshold corresponding on average to the exclusion of 3% of individuals with the highest NETs levels.

For each tested model, the number of times a SNP was found statistically significant at α = 0.05, α = 0.01, and α = 0.001 was used to compute its empirical type I error.

## RESULTS

### Population characteristics

The main characteristics of the FARIVE participants used in this work are presented in **Table 1**. There is approximately 40% of men, 20% of current smokers and individuals are on average 53 years old. The distribution of NETs plasma levels observed in FARIVE is shown in **Figure 1A**. Approximately 16% of exact zeros were observed with a higher proportion among VT cases compared to controls (20% and 10% respectively). To analyse NETs as count data, observed values were multiplied by 1,000. This induced a large overdispersion (mean=78; variance=26,064) leading to the adoption of a Negative Binomial model for analysing such data.

**Table 1.** Main characteristics of the FARIVE study.

|  | **Total N=657** | **VT‡ Cases N=372** | **Controls N=285** |
| --- | --- | --- | --- |
|  | **N (%)** | **N (%)** | **N (%)** |
| **Sex - Men** | 256 (39.0%) | 141 (37.9%) | 115 (40.4%) |
| **Age at sampling** (*Mean ± SD*□) | 53.0 ± 18.8 | 53.3 ± 19.3 | 52.8 ± 18.2 |
| **Smoking status** |  |  |  |
| Current smoker | 128 (19.5%) | 62 (16.7%) | 66 (23.2%) |
| Former smoker / Never | 529 (80.5%) | 310 (83.3%) | 219 (76.8%) |
| **Neutrophil Extracellular Traps levels** |  |  |  |
| All values |  |  |  |
| Mean ± SD | 0.08 ± 0.16 | 0.05 ± 0.11 | 0.12 ± 0.20 |
| Median [Q1;Q3] | 0.03 [4x10<sup>-3</sup>;0.07] | 0.03 [2x10<sup>-3</sup>;0.05] | 0.04 [0.01;0.12] |
| Exact zero | 104 (15.8%) | 75 (20.2%) | 29 (10.2%) |

□ Standard Deviation
‡ Venous Thrombosis

**Figure 1:**
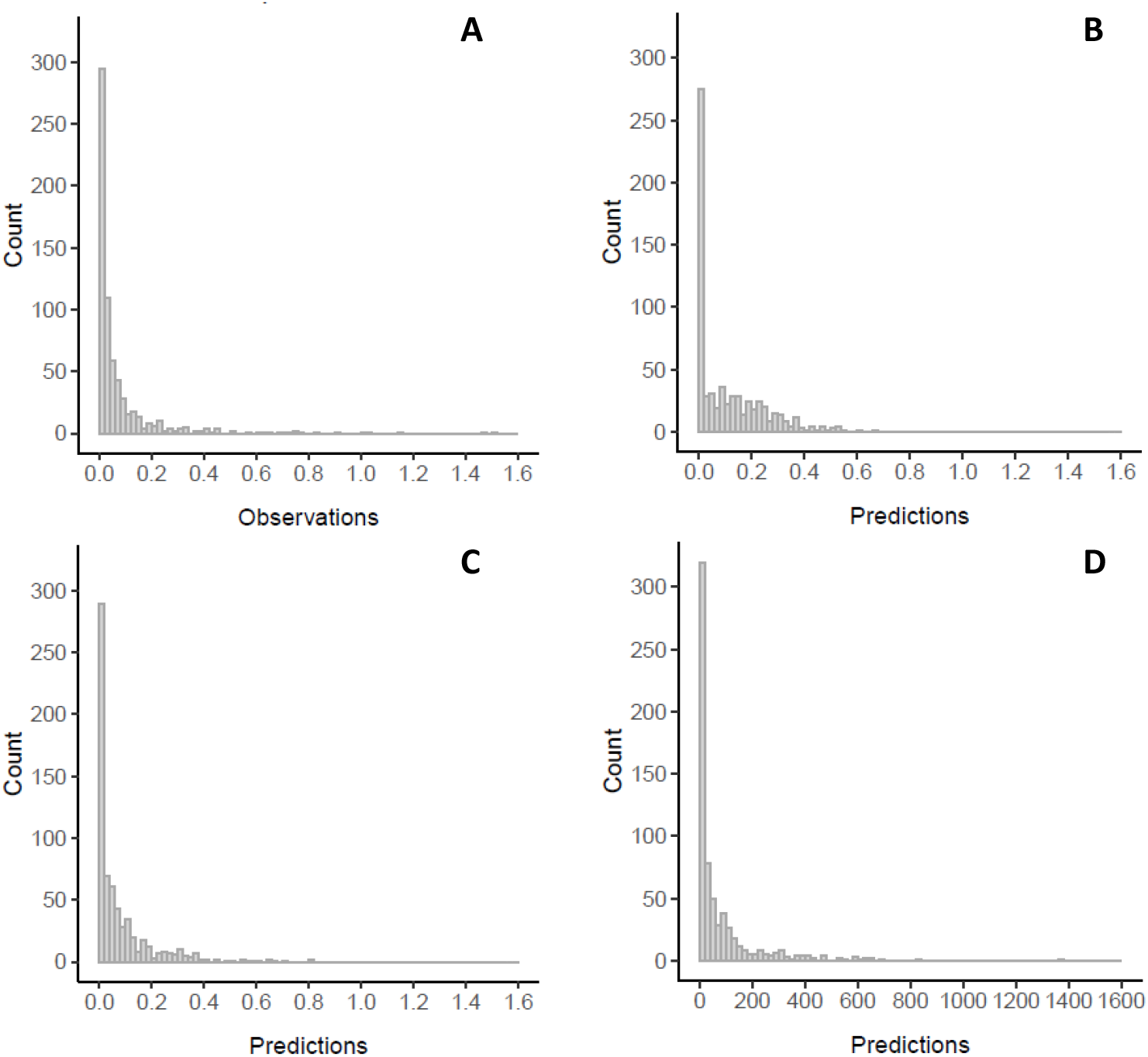
Distribution of NETs plasma levels. This figure presents the distribution of the observed NETs plasma levels (A), predictions from the Tobit model (B), the Compound Poisson-Gamma model (C) and the Negative Binomial model (D).

### Clinical Variables & Goodness of fit

**Table 2** reports the association of clinical covariates with NETs plasma levels in each of the three studied model. The Tobit model assumes a linear association of the covariates on the expected mean of the latent variable, i.e. the true value of NETs. For example, each 10-year increase in age is associated with an increase of 0.005 on the expected mean of the latent variable of NETs plasma levels, given the other covariates are held constant. Regarding the two other models, as a logarithmic link function is used, the association of covariates on the expected mean of NETs is multiplicative. As a consequence, for the Compound Poisson-Gamma model, each 10-year increase of age is associated with an expected mean of NETs plasma levels multiplied by 1.05 (= *e*^0.05^). Similar interpretation holds for the Negative Binomial model that yielded regression parameters very close to those obtained via the Compound Poisson-Gamma models.

**Table 2:** Comparison of regression parameter estimates according to the three models.

|  | **Tobit** | **Compound**<br>**Poisson-Gamma** | **Negative**<br>**Binomial\*** |
| --- | --- | --- | --- |
|  | Beta (SD) | Beta (SD) | Beta (SD) |
| *Covariates* |  |  |  |
| **Age (10y)** | 0.005 (0.004) | 0.05 (0.04) | 0.05 (0.04) |
| **Sex (Males)** | -0.01 (0.02) | -0.08 (0.16) | -0.04 (0.13) |
| **Smoking (Non smokers)** | 0.05 (0.02) | 0.50 (0.19) | 0.52 (0.17) |
| **Status (Controls)** | -0.08 (0.01) | -0.87 (0.15) | -0.87 (0.13) |
| **RMSE\*** | 198.7†<br>[145.5 ; 251.9] | 193.9<br>[180.8 ; 207.1] | 211.5<br>[187.0 ; 236.0] |

\*: For the distribution of NETs multiplied by 1,000. Mean [min-max] over 1,000 bootstrapped samples
†: Negative predictions were censored at zero

**Table 3:**
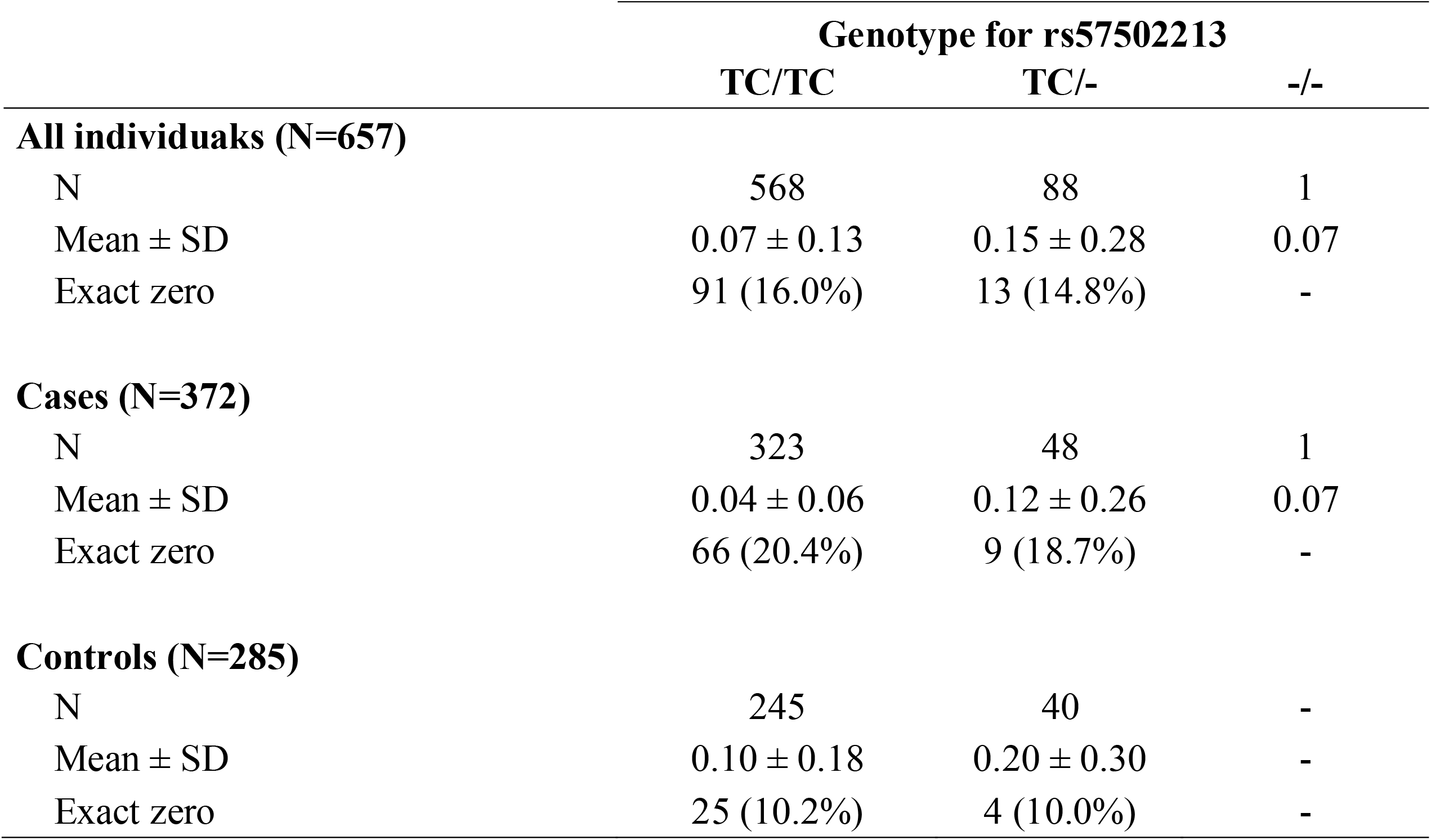
Association of rs57502213 with NETs plasma levels.

|  | Genotype for rs57502213 |  |  |
| --- | --- | --- | --- |
|  | TC/TC | TC/- | -/- |
| **All individuals (N=657)** |  |  |  |
| N | 568 | 88 | 1 |
| Mean ± SD | 0.07 ± 0.13 | 0.15 ± 0.28 | 0.07 |
| Exact zero | 91 (16.0%) | 13 (14.8%) | - |
| **Cases (N=372)** |  |  |  |
| N | 323 | 48 | 1 |
| Mean ± SD | 0.04 ± 0.06 | 0.12 ± 0.26 | 0.07 |
| Exact zero | 66 (20.4%) | 9 (18.7%) | - |
| **Controls (N=285)** |  |  |  |
| N | 245 | 40 | - |
| Mean ± SD | 0.10 ± 0.18 | 0.20 ± 0.30 | - |
| Exact zero | 25 (10.2%) | 4 (10.0%) | - |

RMSEs provided by the three models are shown in **Table 2**. The lowest RMSE was observed for the Compound Poisson-Gamma model while the Negative Binomial model exhibited the highest one. Graphically, the Compound Poisson-Gamma (**Figure 1C**) and the Negative Binomial (**Figure 1D**) showed similar distributions of their predicted values even if the right skewedness was slightly less pronounced for the Compound Poisson-Gamma distribution. These distributions were rather close to that observed for the original NETs data (**Figure 1A**). By contrast, the Tobit distribution (**Figure 1B**) substantially deviated from the original data and looked like a left-truncated Gaussian distribution.

Quantile-Quantile plots of the observed vs predicted values did not visually show obvious deviation from the bisection line, except for high values (above 0.5 in the original NET scale), for the Compound Poisson-Gamma (**Figure 2B**) and the Negative Binomial (**Figure 2C**) models. Conversely, for the Tobit model (**Figure 2A**), the QQplot line deviated from the bisection line from the lower values.

**Figure 2:**
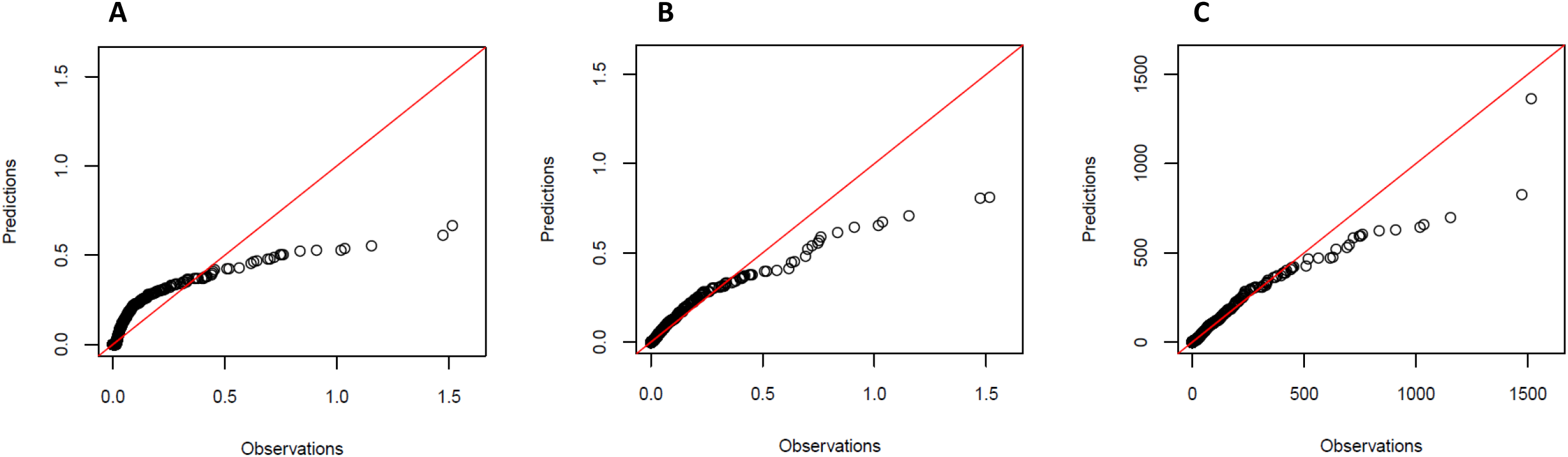
QQplots of observations and predictions from the three models. From Tobit (A), Compound Poisson-Gamma (B) and Negative Binomial (C).

Altogether, these observations suggest that the Compound Poisson-Gamma model seems the most adequate to analyze FARIVE NETs data. Nevertheless, we conducted a GWAS on NETs plasma levels using each of the three models discussed above in order to get additional elements of comparison between these models.

### GWAS analysis on NETs plasma levels

A total of 9,670,724 autosomal SNPs were tested for association with NETs plasma levels using the Tobit, Negative Binomial and Compound Poisson-Gamma models. Quantile-Quantile plots for the observed and expected p-values summarizing the GWAS results for each model are shown in **Figure 3**. While the whole set of association results was compatible with what was expected under the null hypothesis of no genetic association for the Compound Poisson-Gamma model **(Figure 3B**), strong deviations were observed for the Tobit (**Figure 3A**) and Negative Binomial (**Figure 3C**) models. By restricting the GWAS results to SNPs with MAF greater than 5%, inflation was no longer observed for the Tobit model (**Supplementary Figure S1A**, genomic inflation factor λ=0.96) while the Negative Binomial model remained strongly inflated (**Supplementary Figure S1C**, λ=1.46).

**Figure 3:**
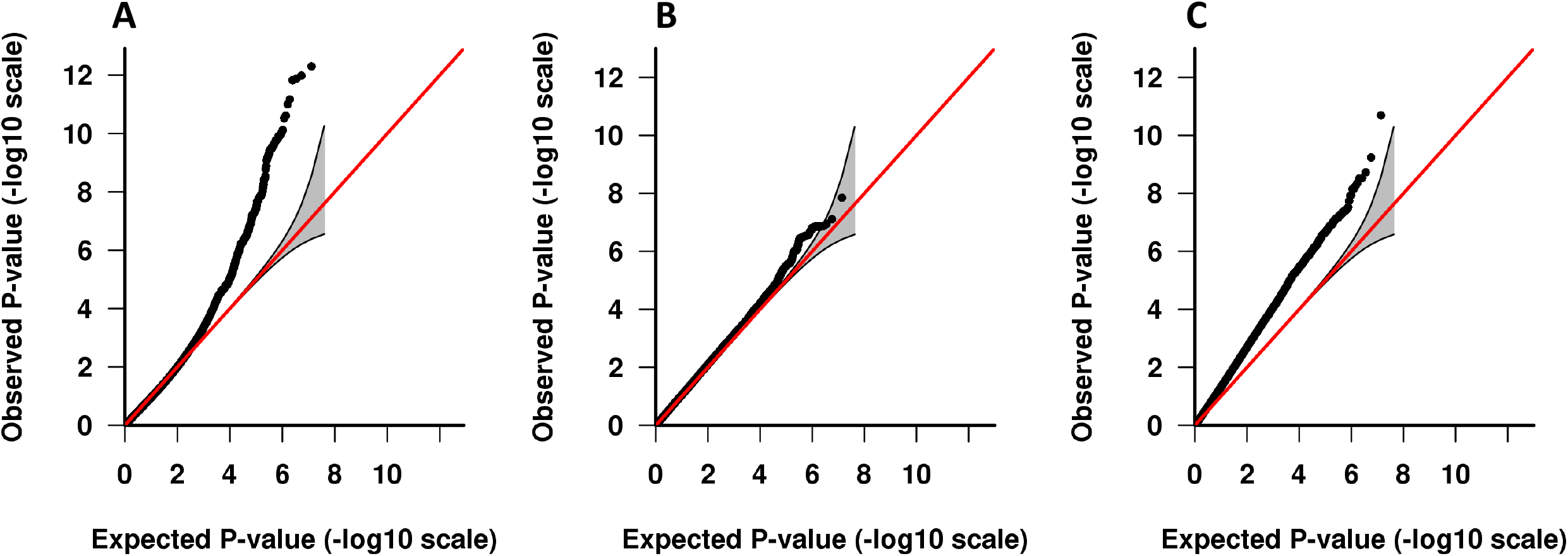
Quantile-Quantile plots from the GWAS results on NETs plasma levels. Tobit (A), Compound Poisson-Gamma (B) and Negative Binomial (C).

To further explore the remaining inflation, we re-ran the GWAS under the Compound Poisson-Gamma and Negative Binomial models after excluding 19 FARIVE participants (∼3%) with NETs plasma levels higher than 0.5. Inflation in the Negative Binomial model was considerably decreased (**Supplementary Figure S2B**) and completely vanished when we additionally restricted the GWAS analysis on SNP with MAF > 5% (**Supplementary Figure S3B**, λ=1.03). Finally, the simulation studies demonstrated that the test of SNP in the Negative Binomial model exhibited inflated type I error for *α* for the three nominal values of alpha considered (**Supplementary Table S1**) when data distribution fit the one here observed for NETs plasma levels in the FARIVE study. Type I error was rather well controlled in absence of extreme values (**Supplementary Table S2**). Of note, these simulations also show that the Compound Poisson-Gamma model generally controls the nominal type I error. All these observations add support for the use of the Compound Poisson-Gamma model for the GWAS analysis of NETs plasma levels.

The corresponding Manhattan plot shown in **Figure 4** revealed one genome-wide significant locus. The lead SNP rs57502213 is a deletion of two nucleotides (TC), mapping to the miR-155 hosting gene (MIR155HG). This variant had a MAF of ∼7%, exhibited a good imputation quality (Rsq=0.92) and its minor allele was associated with a 2.53 fold increase (95% confidence interval [1.85 – 3.47], *P*=1.42×10^-8^) in NETs plasma levels. The average NETs plasma levels were higher in carriers of the deletion of the TC allele than in non-carriers (0.15 vs 0.07). This pattern of association was consistent in VT cases and in controls (**Table 4**). Full GWAS summary statistics will be available on GWAS catalog^49^.

**Figure 4:**
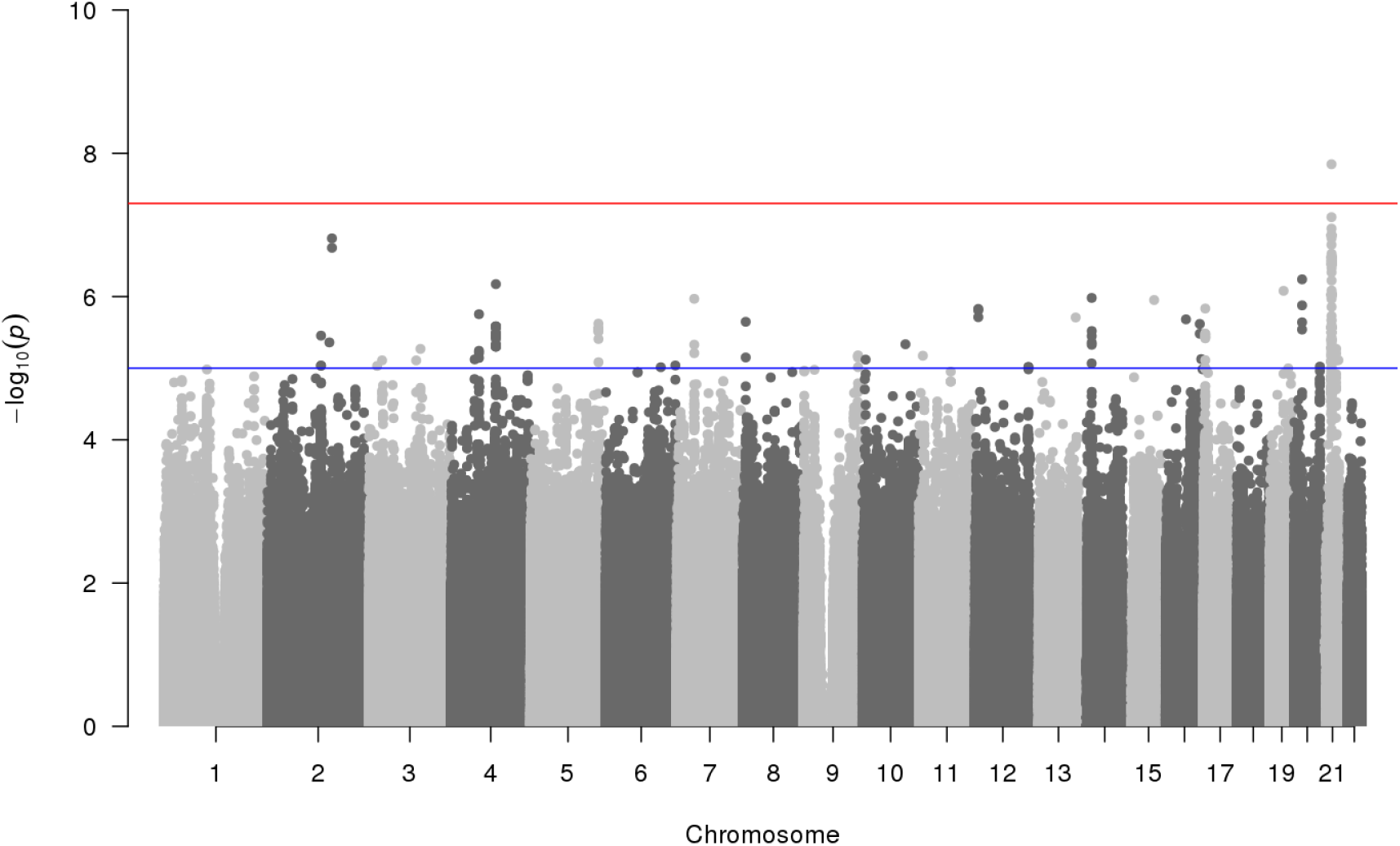
Manhattan plots from the GWAS results of NETs conducted with Compound Poisson-Gamma model. The −*log*_10_ of the p-values are presented according to the position of the associated tested SNP across the genome. The genome wide significant threshold (5×10^-8^) is represented with a red line.

## Discussion

This work was motivated by the search of genetic factors associated with NETs plasma levels exhibiting a semicontinuous distribution. We compared three different modeling strategies, Tobit, Negative Binomial and Compound Poisson-Gamma models, that handle the excess of zero and the asymmetric distribution while allowing the estimation of a single regression parameter for characterizing the association between an explanatory variable and the global mean of the semicontinuous outcome.

Visual inspection showed that both the Negative Binomial and Compound Poisson-Gamma models fit better the observed NETs distribution than the Tobit model. Indeed, the underlying hypothesis of a left-truncated Gaussian distribution with only two parameters makes the Tobit model less-flexible than Compound Poisson-Gamma and Negative Binomial models. RMSE analysis provided further support for the use of Compound Poisson-Gamma model. Of note, the definition of this model matches quite well the biological mechanisms underlying NETs production as it is intuitively reasonable to assume that the number of dead neutrophils follows a Poisson distribution, and that each of these rejects a certain quantity of NETs that would follow a Gamma distribution.

Our GWAS and simulation studies revealed that the Tobit and Negative Binomial models were prone to strong inflation of p-values. While this inflation could be attributable to SNPs with low allele frequency (MAF<5%) for the Tobit model, this inflation was due to both low allele frequency SNPs and extreme positive values of the outcome for the Negative Binomial model. The Compound Poisson-Gamma model was much more robust to these two phenomena.

The Compound Poisson-Gamma based GWAS identified one significant locus on chr21q21.3 associated with NETs plasma levels. This locus maps to miR155HG and the lead SNP was rs57502213, an intronic deletion in miR155HG. While several recent publications have highlighted the role of the associated miRNA in the NETs formation^50–52^, little information is available in public resources about the possible functional impact of rs57502213. This SNP is in moderate linkage disequilibrium (pairwise D’ > 0.50) with several other nearby variants located in *MRPL39, GABPA* and *APP*, the latter having also been reported to be involved in NETs formation^53^. Note that another GWAS on NETs plasma levels has recently been conducted in the Rotterdam study^54^. Despite the large sample size of this study (∼5600 individuals), no significant genome-wide association was detected and the association of our lead SNP did not replicate there (*P*=0.14). However, different kits were used to measure NETs levels in the two studies and recent work has emphasized the need for standardized methods for NETs measurements^55,56^ (https://cdn.ymaws.com/www.isth.org/resource/resmgr/ssc/ssc_subcommittee_project_mar.pdf). Of note, in the Rotterdam study, NETs were analyzed using a log-transformed model suggesting that no zero values (or little) were observed. This contrasts with FARIVE data and could contribute to the heterogeneity of findings between studies. Nevertheless, because of the increasingly recognized role of the chr21q21.3 locus in NETs biology, further works deserve to be conducted to clarify the genetic signal observed in the present study.

To conclude, our work indicates that the modeling strategy for a semicontinuous outcome is crucial, but not straightforward. The choice of the model should take into account the nature of the (biological) process generating zero values, the distribution of the outcome and, especially, the presence of extreme values. The Tobit model with only two parameters is less flexible than Compound Poisson-Gamma and Negative Binomial models and our work suggests that the Compound Poisson-Gamma model, while still marginally used, is more robust than the Negative Binomial model to outliers and low allele frequency making. This make it well suitable for GWAS analysis on semicontinuous trait.

## Data Availability

All data produced in the present study are available upon reasonable request to the authors

## Statements and Declarations

## Acknowledgments

GM benefited from the EUR DPH, a PhD program supported within the framework of the PIA3 (Investment for the future). Project reference 17-EURE-0019.

Statistical analyses benefited from the CBiB computing centre of the University of Bordeaux.

This project was carried out in the framework of the INSERM GOLD Cross-Cutting program (P-EM, D-AT).

## Ethics approval

Research have been performed in accordance with the Declaration of Helsinki. The FARIVE study was approved by the “Comité consultatif de protection des personnes dans la recherche biomedicale” (Project n° 2002-034).

## Consent to participate

Written informed consent to participate was obtained from all FARIVE participants.

---

**Supplementary Figure S1:**
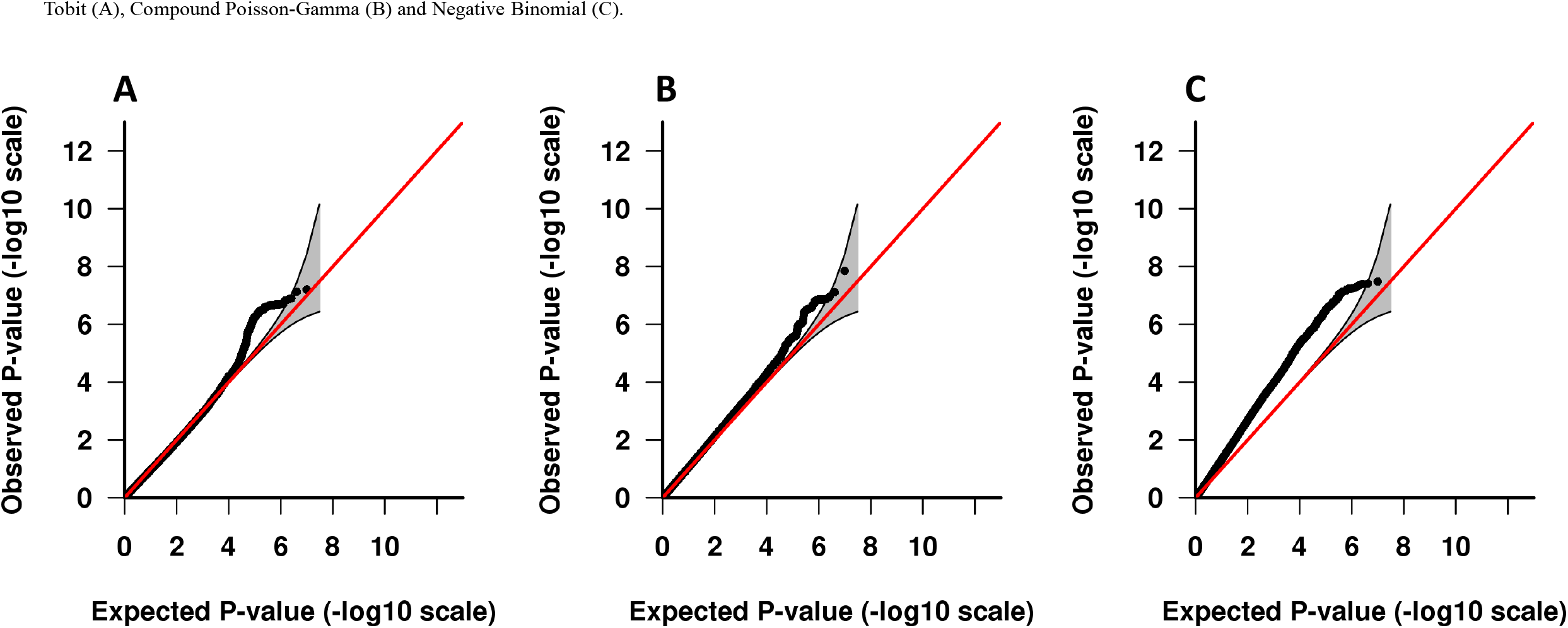
Quantile-Quantile plots from the GWAS results with an allele frequency higher than 5% on NETs plasma levels. Tobit (A), Compound Poisson-Gamma (B) and Negative Binomial (C).

**Supplementary Figure S2:**
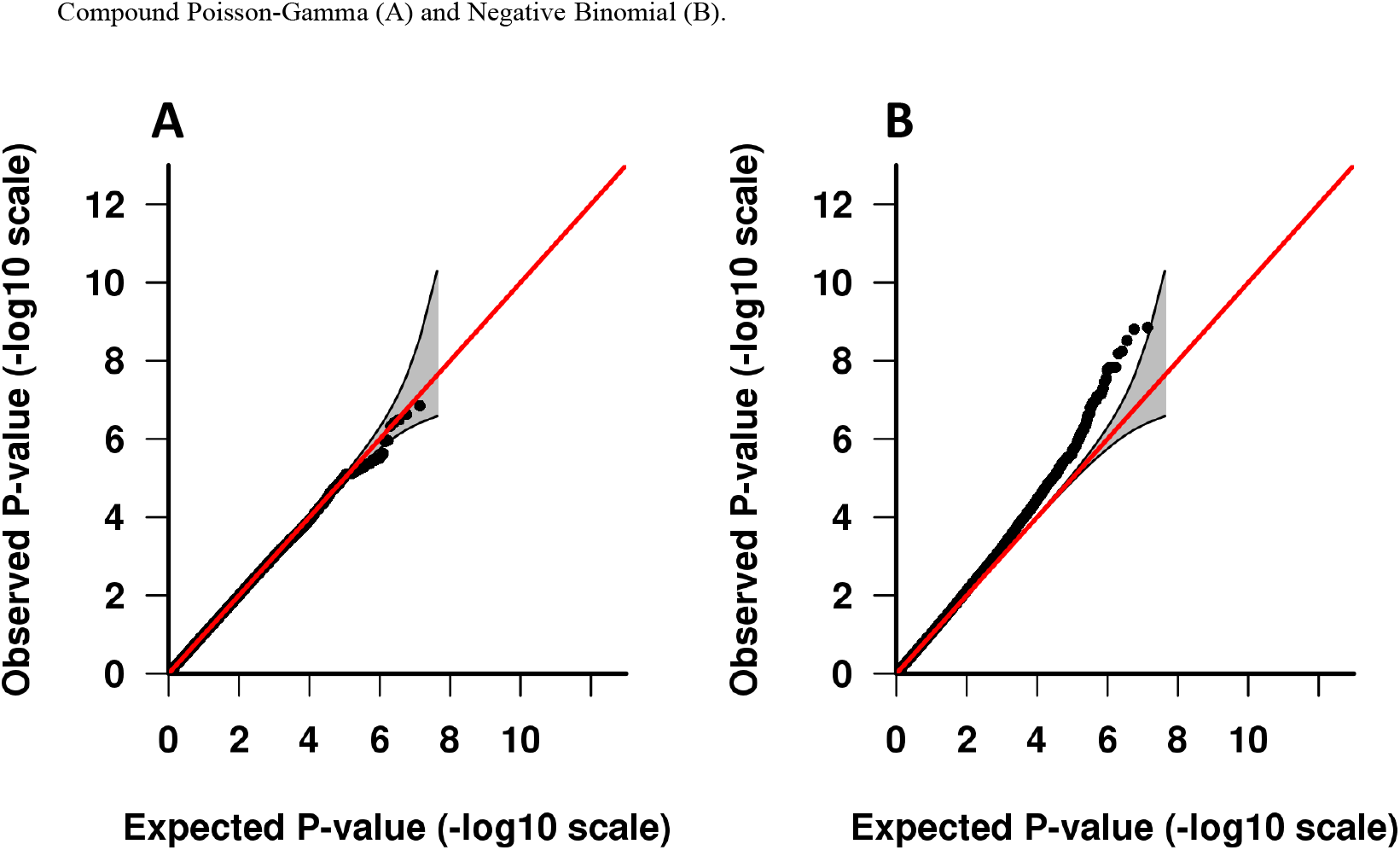
Quantile-Quantile plots from the GWAS results on NETs plasma levels without outliers. Compound Poisson-Gamma (A) and Negative Binomial (B).

**Supplementary Figure S3:**
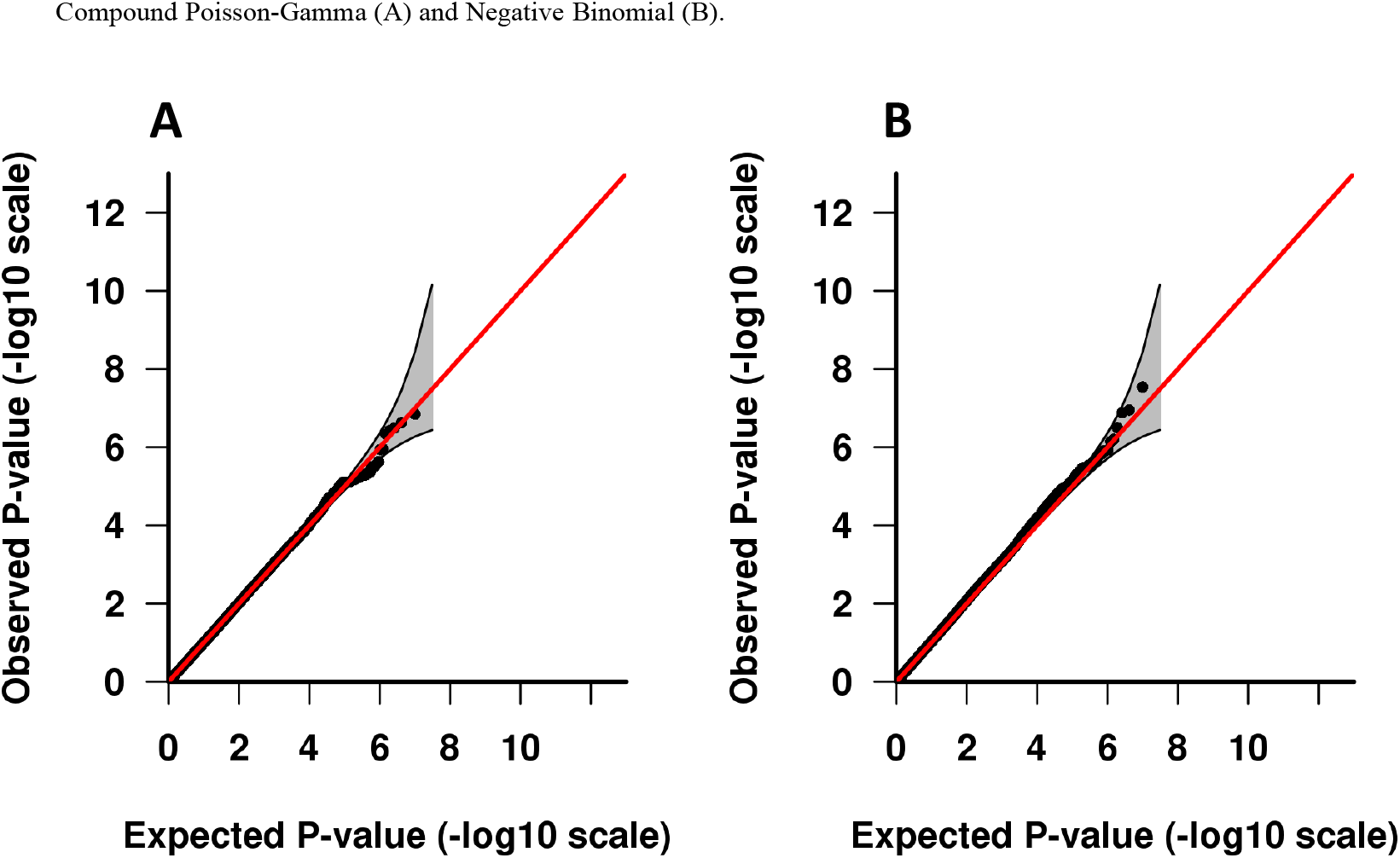
Quantile-Quantile plots from the GWAS results with an allele frequency higher than 5% on NETs plasma levels without outliers. Compound Poisson-Gamma (A) and Negative Binomial (B).

**Supplementary Table S1:** Evaluation of the control of the type I error in 10 000 botstrap samples according to the frequency of the SNP tested with Compound Poisson-Gamma and Negative Binomial models.

| **$\alpha$** | **Compound Poisson-Gamma** |  |  |  | **Negative Binomial** |  |  |  |
| --- | --- | --- | --- | --- | --- | --- | --- | --- |
|  | **MAF 1%** | **MAF 5%** | **MAF 10%** | **MAF 20%** | **MAF 1%** | **MAF 5%** | **MAF 10%** | **MAF 20%** |
| **0.05** | 0.043 | 0.053 | 0.054 | 0.059 | 0.119 | 0.112 | 0.117 | 0.114 |
| **0.01** | 0.007 | 0.010 | 0.013 | 0.013 | 0.039 | 0.036 | 0.038 | 0.038 |
| **0.001** | 0.0004 | 0.0014 | 0.0017 | 0.0011 | 0.0099 | 0.0077 | 0.0084 | 0.0072 |

**Supplementary Table S2:**
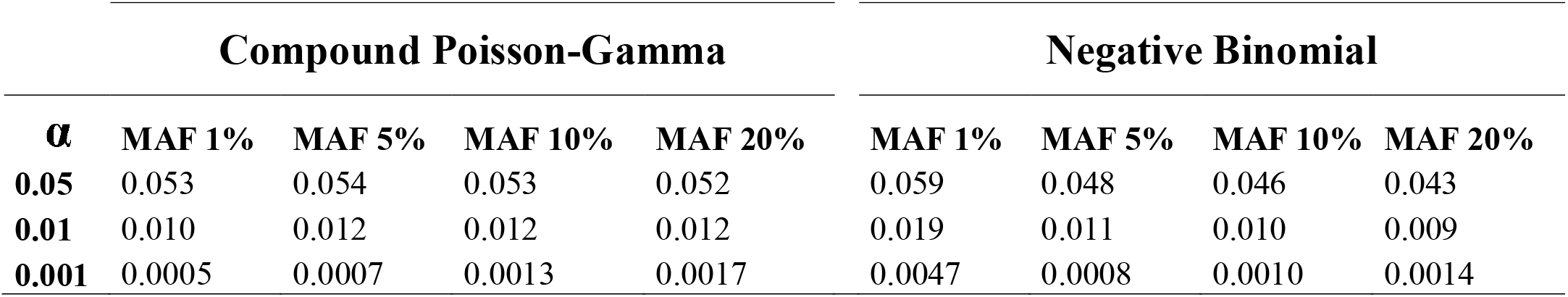
Evaluation of the control of the type I error in 10 000 botstrap samples when outliers are removed, according to the frequency of the SNP tested with Compound Poisson-Gamma and Negative Binomial models.

| **$\alpha$** | **Compound Poisson-Gamma** |  |  |  | **Negative Binomial** |  |  |  |
| --- | --- | --- | --- | --- | --- | --- | --- | --- |
|  | **MAF 1%** | **MAF 5%** | **MAF 10%** | **MAF 20%** | **MAF 1%** | **MAF 5%** | **MAF 10%** | **MAF 20%** |
| **0.05** | 0.053 | 0.054 | 0.053 | 0.052 | 0.059 | 0.048 | 0.046 | 0.043 |
| **0.01** | 0.010 | 0.012 | 0.012 | 0.012 | 0.019 | 0.011 | 0.010 | 0.009 |
| **0.001** | 0.0005 | 0.0007 | 0.0013 | 0.0017 | 0.0047 | 0.0008 | 0.0010 | 0.0014 |

